# Cancer and Post-therapy Cardiotoxicity Risk in Adolescents, Young Adults, and Adults with Down Syndrome

**DOI:** 10.1101/2025.02.21.25322697

**Authors:** Michelle A. Buckman, Anastasiia Vasileva, Charles R. Jedlicka, Hardik Kalra, Mikhail Vasilyev, David S. Dickens, Michael H. Tomasson, Melissa L. Bates

## Abstract

The median life expectancy of people with Down syndrome has increased substantially over the past several decades, from 4 years in 1970 to 53 years in 2010. Despite the recent improvement in survival, there is little data about the prevalence of age-related diseases, including age-related malignancies, and the impact of standard cancer treatments on cardiovascular health. We retrospectively reviewed medical records for age- and sex-matched patients ≥15 years old with and without Down syndrome using the TriNetX platform to identify the prevalence of malignancies and explore cardiovascular outcomes after treatment with anthracyclines. We further stratified the populations into adolescent and young adult (AYA, ages 15-39 years old) and adult (≥40 years old) cohorts, given that treatment recommendations can be different. Down syndrome patients in the AYA cohort were more likely to be diagnosed with acute myeloid (OR 8.9, CI 4.99-15.89, p<0.001) and lymphoid (OR 7.33, CI 4.82-11.15, p<0.001) leukemia. The adult cohort with Down syndrome was more likely to be diagnosed with myelodysplastic syndromes (OR 12.25, CI 6.41-23.42, p<0.001), multiple myeloma (OR 1.66, CI 1.06-2.6, p=0.026), and testicular cancer (OR 2.73, CI 1.32-5.65, p=0.005). Overall, Down syndrome patients (≥15 years old) treated with anthracyclines were more likely to be diagnosed with heart failure (OR 2.14, CI 1.07-4.27, p=0.042). Our study demonstrates adolescents and adults with Down syndrome have a higher predisposition to several malignancies and an increased risk of cardiovascular disease after anthracycline treatment and may require specific screening guidelines to address their unique health risks.

## INTRODUCTION

Since 1950, there has been a four-fold increase in the population prevalence of Down syndrome (DS) in the United States, with a current frequency of 1 in 1,499 live births (1). Since 1970, the median age at death has increased from 4 to 53 years, such that the DS population is both larger and older than in previous decades (2). This increase in life expectancy can be attributed to reduced institutionalization, advancement in surgical treatments of congenital heart disease, and better-established healthcare guidelines, including routine physical and laboratory examinations for children with DS (1, 3, 4)Given the increase in the median life expectancy in DS, it is necessary to understand their susceptibility to age-related co-morbidities and tailor screening and treatment guidelines to meet the needs of this understudied population.

The imbalance in gene dosage from the triplication of human chromosome 21 is associated with accelerated aging (5). This is partly due to the overexpression of oxidative stress-related genes on human chromosome 21, including *SOD1*, *ETS2*, *S100*, and *NIRP1* (6–9). Age-related cancer risk is under-explored. Most studies of cancer prevalence and DS focus only on children or do not stratify the population by age for analysis (10–12). Krieg et al. found no increased risk of cancer in adults with DS (13), but this study focused only on cancers observed in their study population after five years and, therefore, the sample size and the limited 5-year follow-up period may have limited the ability to detect population-level differences, particularly in rarer malignancies. Hasle et al. found a lower-than-expected incidence of breast cancer in individuals with DS, but the small cohort (113 cancer cases overall) limits the ability to evaluate risk except for the most common malignancies (14). The current data are insufficient to develop comprehensive, age-specific cancer screening guidelines for the DS population (15, 16).

Anthracyclines are a class of anti-neoplastic drugs that inhibit DNA synthesis in rapidly dividing cells and are commonly used to treat blood and solid tumor cancers (17). Chronic cardiotoxicity is a serious side effect of anthracycline treatment, and survivors of childhood leukemia with DS are at an increased risk of heart failure (18, 19). However, the effect of anthracyclines on the cardiovascular system of individuals with DS >15 years old is unknown. In the general population, anthracycline-induced cardiovascular toxicity clinically manifests as left ventricular dysfunction, congestive heart failure, cardiomyopathy, cardiac arrhythmias, and/or alterations in cardiac structure (20). We previously found that adults with DS are at a higher risk of hypertension, hypotension, cerebrovascular disease, and ischemic heart disease compared to age- and sex-matched controls and that risk factors used to predict risk in the control population are not necessarily predictive of cardiovascular disease risk in DS (21). Given the overall increased cardiovascular disease risk in DS and the lack of predictive value of typical risk factors, it is critical to independently identify whether they are at an increased risk of chronic cardiotoxicity with anthracycline therapy. Furthermore, adolescent and young adult (AYA) patients are often treated with pediatric-inspired regimens that can include increased exposure to anthracyclines (22), which may alter cardiovascular risk. This highlights the importance of studying the non-pediatric DS population as two cohorts whenever possible – AYA (age 15-39 years) and adult (≥40 years).

This study’s purpose was to comprehensively investigate the prevalence of solid and hematological malignancies in AYA and adult patients with DS by retrospectively analyzing a large medical record database. We further tested the hypothesis that adults with DS who have a history of treatment with anthracyclines will have a higher prevalence of post-treatment cardiovascular disease compared to adults without DS.

## METHODS

### Study design and patient population

A retrospective review of medical records was conducted in January 2025 using the research network of the TriNetX platform, an electronic medical records database consisting of de-identified patient information that can be accessed in aggregate. At the time of the study, the TriNetX platform comprised 117,229,846 adults from 103 healthcare organizations in five countries, mostly in the United States. Data on the TriNetX platform include ICD-10-CM (International Classification of Diseases, 10^th^ Revision, Clinical Modification) diagnosis codes, demographics, medications, lab results, and procedure codes. This study was determined to be exempt from review by the Institutional Review Board at the University of Iowa as data are only reported in aggregate, and identifying personal level data is not available.

Retrospective medical records were reviewed in aggregate for patients ≥15 years old, with the first documented outpatient or inpatient care services as the index event for future follow-up. These patients were sub-grouped into two cohorts: patients with and without DS. The ICD10 code for DS identified individuals with DS. Site-specific cancers and cardiovascular disease types were also determined using ICD-10 diagnosis codes [Supplemental Table 1].

### Data Analysis

The cohorts were matched based on their age at the index event and female sex using 1:1 propensity score matching with the balance cohorts function of TriNetX. We evaluated the incidence of malignant neoplasms occurring in the primary organ systems of both cohorts, followed by restratification of the cohorts into groups aged 15-39 (AYA) and ≥40 (non-AYA) years old. We then explored the incidence of subtypes of hematologic, and solid malignancies recommended for screening and treatment by the American Cancer Society. The TriNetX software automatically eliminated patients who met the index event over 20 years ago. The time window for cancer prevalence started on the day of the first inpatient or outpatient visit after meeting the cohort age criteria, and for cardiovascular disease, the day after the first documented anthracycline treatment, with no designated end date. We further excluded all patients with a neoplasm diagnosis before the index event so that only diagnoses that occurred at or after age 15 were captured. A measure of association analysis, which compares the fraction of patients with the selected neoplastic outcome, was performed using the analysis function of the TriNetX platform. Data are reported as the odds ratio for each neoplasm diagnosis and the 95% confidence intervals. TriNetX automatically reports an outcome as 10 when it involves fewer than 10 patients to protect patient privacy.

Sub-group analyses were performed to evaluate the incidence of cardiovascular diseases in both cohorts after treatment with anthracyclines. The cohorts were matched based on age at the index event, in this case, the anthracycline treatment, and female sex. The time window for each outcome was set to the day after the index event. All patients diagnosed with an outcome before the index event were excluded. The incidence of cardiovascular diseases in the cohort with DS was compared to the group without DS and is reported as the odds ratio and 95% confidence interval. Given the heightened risk of cardiovascular disease we previously reported (21), we performed a within-cohort analysis to evaluate the incidence of cardiovascular diseases in age- and sex-matched control and DS patients, comparing those with neoplasms and anthracyclines to those without a record of any neoplasm or anthracycline medication. The index event for the cohort with neoplasms was when they had a record of anthracycline treatment, while that of the other group was when they had a record of an outpatient/inpatient visit.

### Statistics

Statistical significance was set *a priori* as p < 0.05. All statistical analyses were conducted using the analytics feature in the TriNetX research network. The TriNetX software calculated T-tests of demographic variables and the odds ratios and 95% confidence intervals for outcomes using the measure of association analyses feature. Figures and all other statistical analyses for significance were generated with the GraphPad Prism 10.0 software package.

## RESULTS

### Overall Cohort characteristics

A total of 40,371,881 patients with outpatient or inpatient visits were identified, and a flowchart of the analysis is outlined in Figure 1. Patient demographics, co-morbidities, and lifestyle factors are outlined in Table 1. The age at index (37.1 ± 16.0 years) and female sex (2,600, 51.7%) were not different between groups. In the controls, 2,182 (43.4%) were male, while the cohort with DS consisted of 2,188 (43.5%) male patients. Comorbidities investigated in the controls and the DS cohort include congenital malformations of the circulatory system (73, 1.5% and 930, 18.5%), diabetes (300, 6.0% and 498, 9.9%), pre-existing hypotension (87, 1.7% and 372, 7.4%), and pre-existing hypertension (694, 13.8% and 702, 14.0%). Also, lifestyle factors such as alcohol use disorders (115, 2.3% and 25, 0.5%) and tobacco use (115, 2.3% and 30, 0.6%) were less common in the cohort with DS.

**Figure 1.**
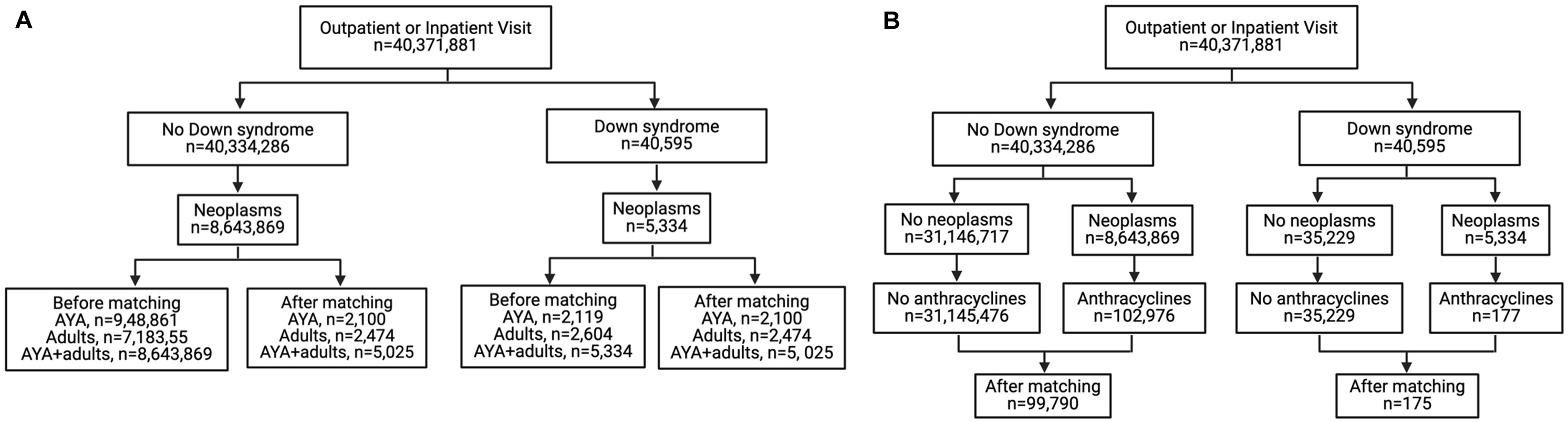
Flow diagram for TriNetX analyses. AYA, adolescent and young adult. Created with BioRender.com

**Table 1.** Baseline characteristics of patients with neoplasms before and after matching.

|  | Before matching |  |  | After matching |  |  |
| --- | --- | --- | --- | --- | --- | --- |
|  | Controls<br>n=8,643,869 | DS<br>n=5,334 | p-value | Controls<br>n=5,025 | DS<br>n=5,025 | p-value |
| **Age, years** |  |  |  |  |  |  |
| †Age at index | 54.7 ± 16.3 | 37.1 ± 16.0 | **< 0.001** | 37.1 ± 16.0 | 37.1 ± 16.0 | 1.000 |
| Current age | 62.0 ± 16.5 | 44.6 ± 16.8 | **< 0.001** | 44.7 ± 16.7 | 44.6 ± 16.8 | 0.712 |
| **Sex** |  |  |  |  |  |  |
| †Female | 4,550,445 (55.7%) | 2,600 (51.7%) | **< 0.001** | 2,600 (51.7%) | 2,600 (51.7%) | 1.000 |
| Male | 3,261,730 (39.9%) | 2,188 (43.5%) | **< 0.001** | 2,182 (43.4%) | 2,188 (43.5%) | 0.904 |
| Unknown sex | 360,916 (4.4%) | 237 (4.7%) | 0.300 | 243 (4.8%) | 237 (4.7%) | 0.779 |
| **Race** |  |  |  |  |  |  |
| White | 5,594,554 (68.5%) | 3,566 (71.0%) | **< 0.001** | 3,311 (65.9%) | 3,566 (71.0%) | **< 0.001** |
| Unknown race | 1,163,321 (14.2%) | 642 (12.8%) | **0.003** | 787 (15.7%) | 642 (12.8%) | **< 0.001** |
| Black or African American | 885,377 (10.8%) | 478 (9.5%) | **0.003** | 568 (11.3%) | 478 (9.5%) | **0.003** |
| Other Race | 223,761 (2.7%) | 190 (3.8%) | **< 0.001** | 170 (3.4%) | 190 (3.8%) | 0.283 |
| Asian | 259,488 (3.2%) | 131 (2.6%) | **0.022** | 156 (3.1%) | 131 (2.6%) | 0.134 |
| Native Hawaiian or other Pacific Islander | 27,614 (0.3%) | 14 (0.3%) | 0.469 | 16 (0.3%) | 14 (0.3%) | 0.715 |
| American Indian or Alaska Native | 18,976 (0.2%) | 10 (0.2%) | 0.625 | 17 (0.3%) | 10 (0.2%) | 0.177 |
| **Ethnicity** |  |  |  |  |  |  |
| Not Hispanic or Latino | 5,781,038 (70.7%) | 3,415 (68.0%) | **< 0.001** | 3,468 (69.0%) | 3,415 (68.0%) | 0.255 |
| Hispanic or Latino | 618,432 (7.6%) | 633 (12.6%) | **< 0.001** | 513 (10.2%) | 633 (12.6%) | **< 0.001** |
| Unknown Ethnicity | 1,773,621 (21.7%) | 977 (19.4%) | **< 0.001** | 1,044 (20.8%) | 977 (19.4%) | 0.095 |
| **Comorbidities and lifestyle factors** |  |  |  |  |  |  |
| Congenital malformations of the circulatory system | 84,154 (1.0%) | 930 (18.5%) | **< 0.001** | 73 (1.5%) | 930 (18.5%) | **< 0.001** |
| Diabetes mellitus | 919,234 (11.2%) | 498 (9.9%) | **0.003** | 300 (6.0%) | 498 (9.9%) | **< 0.001** |
| Hypotension | 181,276 (2.2%) | 372 (7.4%) | **< 0.001** | 87 (1.7%) | 372 (7.4%) | **< 0.001** |
| Hypertensive diseases | 2,190,297 (26.8%) | 702 (14.0%) | **< 0.001** | 694 (13.8%) | 702 (14%) | 0.818 |
| Overweight and obesity | 1,016,185 (12.4%) | 1,143 (22.7%) | **< 0.001** | 551 (11.0%) | 1,143 (22.7%) | **< 0.001** |
| Alcohol use disorders | 198,956 (2.4%) | 25 (0.5%) | **< 0.001** | 89 (1.8%) | 25 (0.5%) | **< 0.001** |
| Tobacco use | 199,476 (2.4%) | 30 (0.6%) | **< 0.001** | 115 (2.3%) | 30 (0.6%) | **< 0.001** |

Data are represented as mean ± SD or n (%). DS, Down syndrome. All significant values are indicated in bold, p<0.05. †Demographic parameters used to match groups.

### Cancer incidence

After matching, all individuals with DS ≥15 years old were three times more likely to be diagnosed with malignancies of the lymphoid, hematopoietic, and related tissues (OR 3.09, CI 2.65-3.61, p<0.001, Figure 2 and Supplemental Table 2). The likelihood of a diagnosis of any malignancy was not different between groups (OR 1.02, CI 0.93-1.11, p=0.753), and patients with DS were less likely to be diagnosed with any solid malignancy than the control group (OR 0.63, CI 0.57-0.7, p<0.001). However, this decreased risk was not universally applicable to all solid tumor malignancies. (Supplemental Table 2).

**Figure 2.**
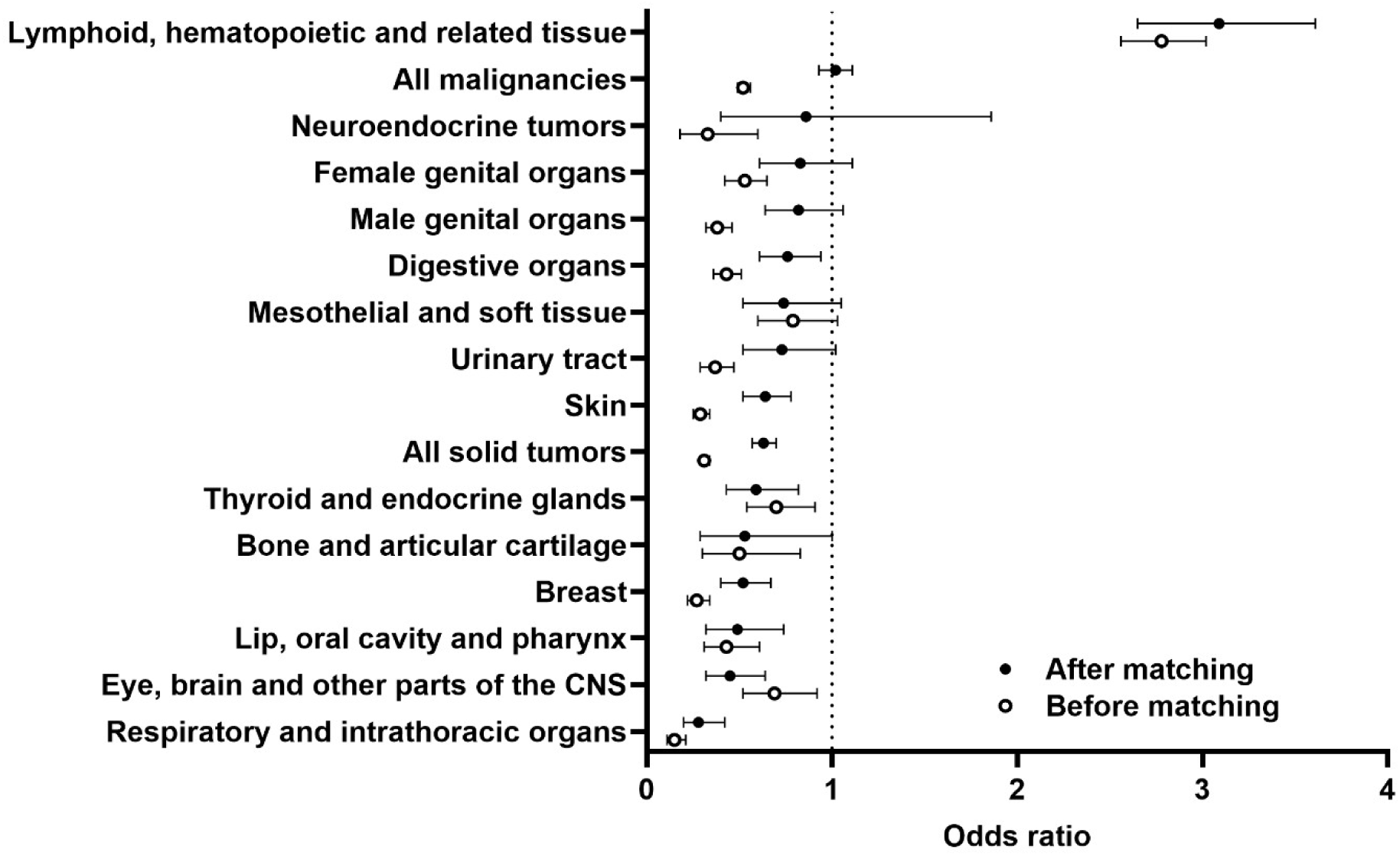
Cancer incidence in the primary organ systems before and after propensity score matching in individuals with and without DS, ≥15 years old. The data are represented as odds ratios with 95% confidence intervals comparing adults with DS to the control cohort. CNS, central nervous system.

Notably, in the matched cohorts, AYA DS patients were less likely to be diagnosed with Hodgkin lymphoma (OR 0.44, CI 0.22-0.89, p=0.028), but were at an increased risk for acute lymphoid and myeloid and chronic myeloid leukemias. [Table 2]. Surprisingly, adults ≥40 years were more likely to be diagnosed with myelodysplastic syndromes (OR 12.25, CI 6.41-23.42, p<0.001) and myeloma (OR 1.66, CI 1.06-2.6, p=0.026) and had an increased risk of all leukemias but not lymphoma. Testicular cancer (OR 2.73, CI 1.32-5.65, p=0.005) was more likely to be diagnosed in individuals 40 years and older with DS, but they were less likely to be diagnosed with breast (OR 0.48, CI 0.36-0.63, p<0.001), and prostate (OR 0.35, CI 0.24-0.53, p<0.001) cancer. [Table 3]. All individuals ≥15 years of age had a lower incidence of cervical (OR 0.52, CI 0.29-0.96, p=0.044) and lung (OR 0.32, CI 0.2-0.51, p<0.001) cancers, likely driven by decreased risk in patients ≥40 years old.

**Table 2.**
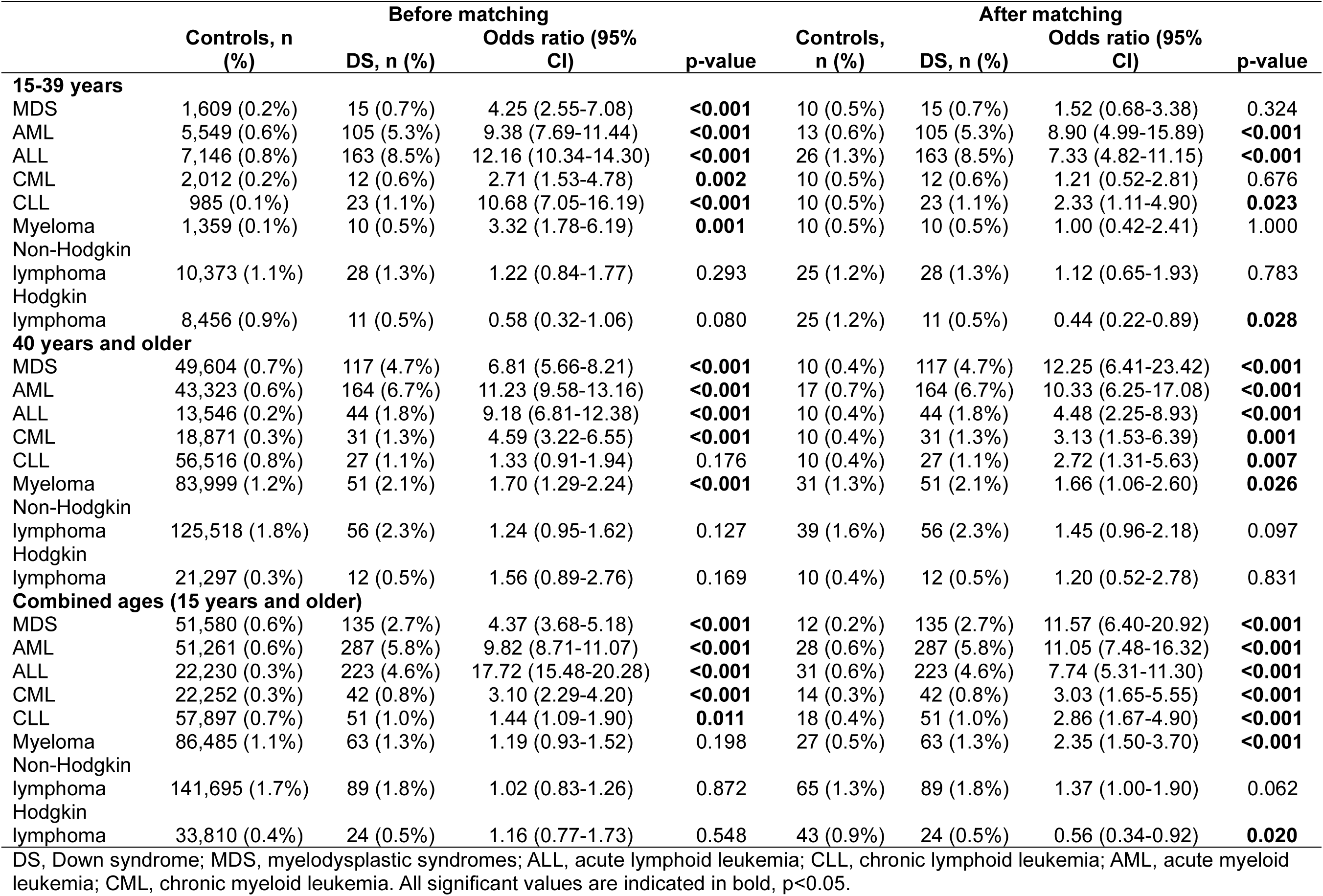
The incidence of blood cancers across the lifespan of individuals with DS compared to those without DS.

|  | Before matching |  |  |  | After matching |  |  |  |
| --- | --- | --- | --- | --- | --- | --- | --- | --- |
|  | Controls, n (%) | DS, n (%) | Odds ratio (95% CI) | p-value | Controls, n (%) | DS, n (%) | Odds ratio (95% CI) | p-value |
| **15-39 years** |  |  |  |  |  |  |  |  |
| MDS | 1,609 (0.2%) | 15 (0.7%) | 4.25 (2.55-7.08) | **<0.001** | 10 (0.5%) | 15 (0.7%) | 1.52 (0.68-3.38) | 0.324 |
| AML | 5,549 (0.6%) | 105 (5.3%) | 9.38 (7.69-11.44) | **<0.001** | 13 (0.6%) | 105 (5.3%) | 8.90 (4.99-15.89) | **<0.001** |
| ALL | 7,146 (0.8%) | 163 (8.5%) | 12.16 (10.34-14.30) | **<0.001** | 26 (1.3%) | 163 (8.5%) | 7.33 (4.82-11.15) | **<0.001** |
| CML | 2,012 (0.2%) | 12 (0.6%) | 2.71 (1.53-4.78) | **0.002** | 10 (0.5%) | 12 (0.6%) | 1.21 (0.52-2.81) | 0.676 |
| CLL | 985 (0.1%) | 23 (1.1%) | 10.68 (7.05-16.19) | **<0.001** | 10 (0.5%) | 23 (1.1%) | 2.33 (1.11-4.90) | **0.023** |
| Myeloma | 1,359 (0.1%) | 10 (0.5%) | 3.32 (1.78-6.19) | **0.001** | 10 (0.5%) | 10 (0.5%) | 1.00 (0.42-2.41) | 1.000 |
| Non-Hodgkin lymphoma | 10,373 (1.1%) | 28 (1.3%) | 1.22 (0.84-1.77) | 0.293 | 25 (1.2%) | 28 (1.3%) | 1.12 (0.65-1.93) | 0.783 |
| Hodgkin lymphoma | 8,456 (0.9%) | 11 (0.5%) | 0.58 (0.32-1.06) | 0.080 | 25 (1.2%) | 11 (0.5%) | 0.44 (0.22-0.89) | **0.028** |
| **40 years and older** |  |  |  |  |  |  |  |  |
| MDS | 49,604 (0.7%) | 117 (4.7%) | 6.81 (5.66-8.21) | **<0.001** | 10 (0.4%) | 117 (4.7%) | 12.25 (6.41-23.42) | **<0.001** |
| AML | 43,323 (0.6%) | 164 (6.7%) | 11.23 (9.58-13.16) | **<0.001** | 17 (0.7%) | 164 (6.7%) | 10.33 (6.25-17.08) | **<0.001** |
| ALL | 13,546 (0.2%) | 44 (1.8%) | 9.18 (6.81-12.38) | **<0.001** | 10 (0.4%) | 44 (1.8%) | 4.48 (2.25-8.93) | **<0.001** |
| CML | 18,871 (0.3%) | 31 (1.3%) | 4.59 (3.22-6.55) | **<0.001** | 10 (0.4%) | 31 (1.3%) | 3.13 (1.53-6.39) | **0.001** |
| CLL | 56,516 (0.8%) | 27 (1.1%) | 1.33 (0.91-1.94) | 0.176 | 10 (0.4%) | 27 (1.1%) | 2.72 (1.31-5.63) | **0.007** |
| Myeloma | 83,999 (1.2%) | 51 (2.1%) | 1.70 (1.29-2.24) | **<0.001** | 31 (1.3%) | 51 (2.1%) | 1.66 (1.06-2.60) | **0.026** |
| Non-Hodgkin lymphoma | 125,518 (1.8%) | 56 (2.3%) | 1.24 (0.95-1.62) | 0.127 | 39 (1.6%) | 56 (2.3%) | 1.45 (0.96-2.18) | 0.097 |
| Hodgkin lymphoma | 21,297 (0.3%) | 12 (0.5%) | 1.56 (0.89-2.76) | 0.169 | 10 (0.4%) | 12 (0.5%) | 1.20 (0.52-2.78) | 0.831 |
| **Combined ages (15 years and older)** |  |  |  |  |  |  |  |  |
| MDS | 51,580 (0.6%) | 135 (2.7%) | 4.37 (3.68-5.18) | **<0.001** | 12 (0.2%) | 135 (2.7%) | 11.57 (6.40-20.92) | **<0.001** |
| AML | 51,261 (0.6%) | 287 (5.8%) | 9.82 (8.71-11.07) | **<0.001** | 28 (0.6%) | 287 (5.8%) | 11.05 (7.48-16.32) | **<0.001** |
| ALL | 22,230 (0.3%) | 223 (4.6%) | 17.72 (15.48-20.28) | **<0.001** | 31 (0.6%) | 223 (4.6%) | 7.74 (5.31-11.30) | **<0.001** |
| CML | 22,252 (0.3%) | 42 (0.8%) | 3.10 (2.29-4.20) | **<0.001** | 14 (0.3%) | 42 (0.8%) | 3.03 (1.65-5.55) | **<0.001** |
| CLL | 57,897 (0.7%) | 51 (1.0%) | 1.44 (1.09-1.90) | **0.011** | 18 (0.4%) | 51 (1.0%) | 2.86 (1.67-4.90) | **<0.001** |
| Myeloma | 86,485 (1.1%) | 63 (1.3%) | 1.19 (0.93-1.52) | 0.198 | 27 (0.5%) | 63 (1.3%) | 2.35 (1.50-3.70) | **<0.001** |
| Non-Hodgkin lymphoma | 141,695 (1.7%) | 89 (1.8%) | 1.02 (0.83-1.26) | 0.872 | 65 (1.3%) | 89 (1.8%) | 1.37 (1.00-1.90) | 0.062 |
| Hodgkin lymphoma | 33,810 (0.4%) | 24 (0.5%) | 1.16 (0.77-1.73) | 0.548 | 43 (0.9%) | 24 (0.5%) | 0.56 (0.34-0.92) | **0.020** |

DS, Down syndrome; MDS, myelodysplastic syndromes; ALL, acute lymphoid leukemia; CLL, chronic lymphoid leukemia; AML, acute myeloid leukemia; CML, chronic myeloid leukemia. All significant values are indicated in bold, $p < 0.05$ .

**Table 3.** The incidence of solid tumors in individuals with DS compared to individuals without DS grouped into AYA and non-AYA patients.

|  | Before matching |  |  |  | After matching |  |  |  |
| --- | --- | --- | --- | --- | --- | --- | --- | --- |
|  | Controls, n (%) | DS, n (%) | Odds ratio (95% CI) | p-value | Controls, n (%) | DS, n (%) | Odds ratio (95% CI) | p-value |
| **15-39 years** |  |  |  |  |  |  |  |  |
| Testis | 7,805 (0.8%) | 32 (1.5%) | 1.86 (1.31-2.63) | **0.002** | 26 (1.2%) | 32 (1.5%) | 1.23 (0.73-2.08) | 0.509 |
| Endometrium | 2,044 (0.2%) | 10 (0.5%) | 2.20 (1.18-4.11) | **0.029** | 10 (0.5%) | 10 (0.5%) | 1.00 (0.42-2.41) | 1.000 |
| Colon | 3,833 (0.4%) | 10 (0.5%) | 1.17 (0.63-2.18) | 0.603 | 10 (0.5%) | 10 (0.5%) | 1.00 (0.42-2.41) | 1.000 |
| Cervix | 4,725 (0.5%) | 10 (0.5%) | 0.95 (0.51-1.77) | 1.000 | 10 (0.5%) | 10 (0.5%) | 1.00 (0.42-2.41) | 1.000 |
| Breast | 10,578 (1.1%) | 10 (0.5%) | 0.42 (0.23-0.79) | **0.003** | 17 (0.8%) | 10 (0.5%) | 0.59 (0.27-1.28) | 0.246 |
| Prostate | 705 (0.1%) | 10 (0.5%) | 6.40 (3.42-11.96) | **<0.001** | 10 (0.5%) | 10 (0.5%) | 1.00 (0.42-2.41) | 1.000 |
| Lung | 2,749 (0.3%) | 10 (0.5%) | 1.64 (0.88-3.05) | 0.149 | 10 (0.5%) | 10 (0.5%) | 1.00 (0.42-2.41) | 1.000 |
| **40 years and older** |  |  |  |  |  |  |  |  |
| Testis | 12,688 (0.2%) | 27 (1.1%) | 5.98 (4.09-8.74) | **<0.001** | 10 (0.4%) | 27 (1.1%) | 2.73 (1.32-5.65) | **0.005** |
| Endometrium | 102,365 (1.5%) | 25 (1.0%) | 0.67 (0.45-1.00) | 0.058 | 25 (1.0%) | 25 (1.0%) | 1.00 (0.57-1.75) | 1.000 |
| Colon | 181,306 (2.6%) | 36 (1.5%) | 0.54 (0.39-0.76) | **<0.001** | 49 (2.0%) | 36 (1.5%) | 0.73 (0.47-1.13) | 0.189 |
| Cervix | 44,072 (0.6%) | 11 (0.4%) | 0.69 (0.38-1.25) | 0.268 | 18 (0.7%) | 11 (0.4%) | 0.61 (0.29-1.29) | 0.264 |
| Breast | 506,879 (7.4%) | 76 (3.1%) | 0.40 (0.32-0.50) | **<0.001** | 154 (6.3%) | 76 (3.1%) | 0.48 (0.36-0.63) | **<0.001** |
| Prostate | 439,145 (6.4%) | 33 (1.3%) | 0.20 (0.14-0.28) | **<0.001** | 91 (3.7%) | 33 (1.3%) | 0.35 (0.24-0.53) | **<0.001** |
| Lung | 305,229 (4.5%) | 20 (0.8%) | 0.18 (0.11-0.27) | **<0.001** | 78 (3.2%) | 20 (0.8%) | 0.25 (0.15-0.41) | **<0.001** |
| **Combined ages (15 years and older)** |  |  |  |  |  |  |  |  |
| Testis | 25,120 (0.3%) | 82 (1.6%) | 5.38 (4.33-6.70) | **<0.001** | 32 (0.6%) | 82 (1.6%) | 2.59 (1.72-3.9) | **<0.001** |
| Endometrium | 106,735 (1.3%) | 29 (0.6%) | 0.44 (0.31-0.63) | **<0.001** | 28 (0.6%) | 29 (0.6%) | 1.04 (0.62-1.74) | 0.895 |
| Colon | 189,012 (2.3%) | 49 (1.0%) | 0.42 (0.31-0.55) | **<0.001** | 59 (1.2%) | 49 (1.0%) | 0.83 (0.57-1.21) | 0.384 |
| Cervix | 55,479 (0.7%) | 17 (0.3%) | 0.50 (0.31-0.80) | **0.004** | 32 (0.6%) | 17 (0.3%) | 0.53 (0.29-0.96) | **0.044** |
| Breast | 532,575 (6.5%) | 94 (1.9%) | 0.27 (0.22-0.34) | **<0.001** | 179 (3.6%) | 94 (1.9%) | 0.52 (0.40-0.67) | **<0.001** |
| Prostate | 439,537 (5.4%) | 38 (0.8%) | 0.13 (0.10-0.18) | **<0.001** | 98 (2.0%) | 38 (0.8%) | 0.38 (0.26-0.56) | **<0.001** |
| Lung | 309,244 (3.8%) | 24 (0.5%) | 0.12 (0.08-0.18) | **<0.001** | 74 (1.5%) | 24 (0.5%) | 0.32 (0.20-0.51) | **<0.001** |

DS, Down syndrome; AYA, adolescent and young adults. All significant values are indicated in bold, $p < 0.05$ .

### Anthracycline cardiotoxicity

Because of the limited sample size, the incidence of cardiovascular disease after treatment with anthracycline medications was only evaluated for matched individuals ≥15 years old with and without DS. The average available follow-up period was three years. The DS group had an increased rate of ischemic heart diseases (OR 2.3, CI 1.05-5.05, p=0.039), diseases of arteries, arterioles, and capillaries (OR 2.28, CI 1.15-4.51, p=0.20), and heart failure (OR 2.14, CI 1.07-4.27, 0.042). [Supplemental Table 4, Figure 3].

**Figure 3.**
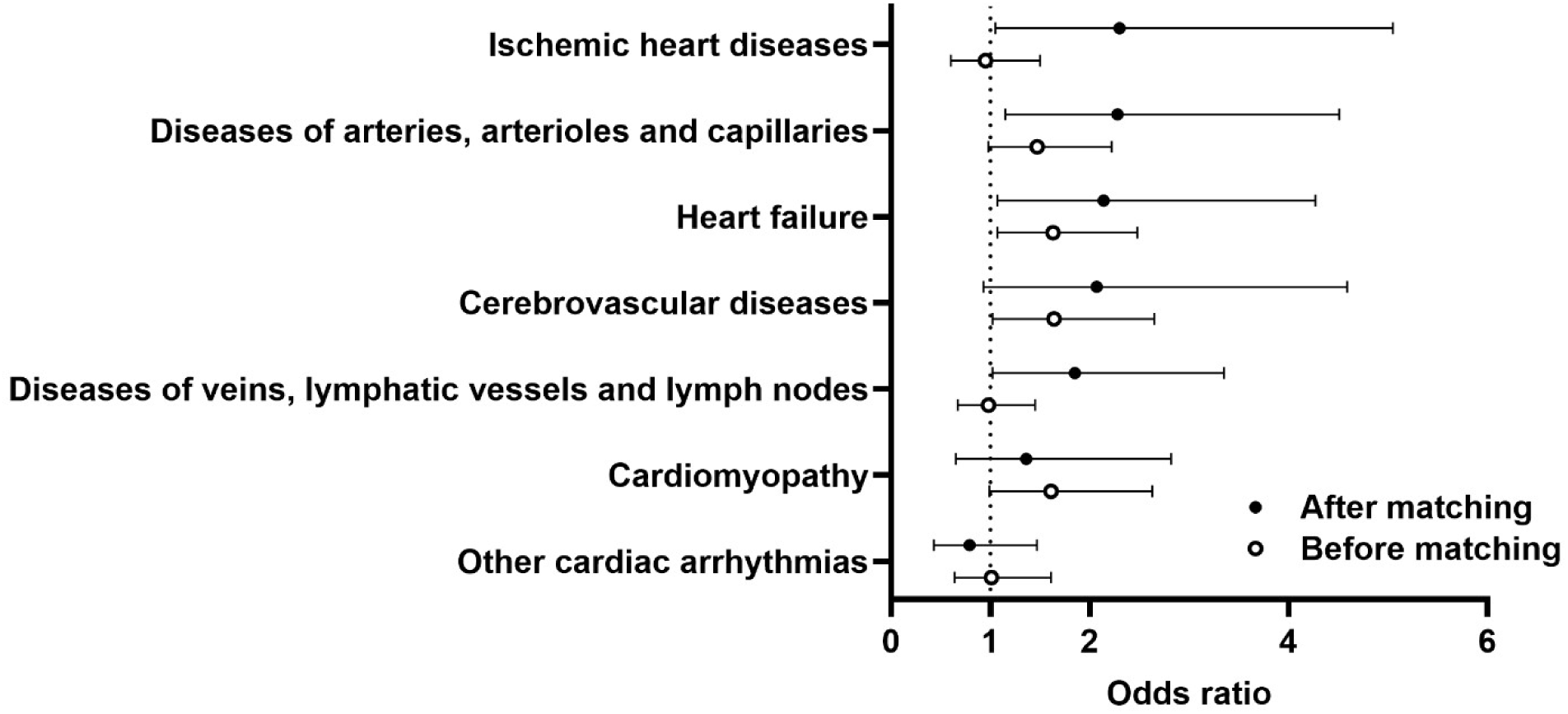
The incidence of cardiovascular diseases in patients with and without DS aged ≥15 years old after treatment with anthracyclines. Data are represented as the odds ratios comparing adults with DS to the control group with 95% confidence intervals.

## DISCUSSION

We conducted a medical records review to assess the occurrence of hematological and solid malignancies in a large cohort of AYA and adults with DS. We found that individuals with DS are three times more likely to be diagnosed with malignancies of the lymphoid, hematopoietic, and related tissues compared to age and sex-matched controls, such that the risk of blood cancer is not unique to the pediatric period. They also have a similar incidence of all malignancies but a lower or similar incidence of some solid tumors. We also report for the first time that their risk of myelodysplastic syndromes and multiple myeloma significantly increases after 40 years of age. Other heme malignancies, such as lymphoid and myeloid leukemia, are commonly diagnosed in both the AYA and non-AYA cohorts with DS. The incidence of ischemic heart diseases, heart failure, and vascular diseases in individuals with DS and anthracycline medications is higher compared to the control group.

### Hematological malignancies

Altered hematopoiesis is one of the health conditions associated with DS, and we previously reported an increased risk of anemia in patients with DS (23). It is well-established that children with DS have a significantly increased risk of developing leukemia compared to children without DS (24). This has been associated with *GATA1* mutations, which primarily drive myeloid leukemia of DS (25). Also, the overexpression of genes, including *DYRK1A*, *RCAN1*, *RUNX1*, *ERG*, and *HMGN1* on human chromosome 21, has been implicated in DS leukemogenesis (26–28). Like children with DS, our study shows that the risk of leukemia persists in adults. Kreig et al. found no significant difference in the risk of leukemia in adults ≥18 years old with DS compared to adults without DS (13). However, Hasle et al. found a high risk of leukemia in young adults aged 10-29 years, with only three cases of leukemia in individuals >30 years old (10). The variability in the results may be due to the larger sample size in our study, which provided access to a broader population of adults with Down syndrome. Additional research is needed to investigate the mechanisms of leukemogenesis in AYA and older adult patients with DS, which will improve treatment options for this emerging population. It is essential to acknowledge that some cases of leukemia in our adult population could be due to relapses from childhood leukemia since patients with DS and acute lymphoid leukemia have a higher risk of relapse compared to patients without DS (29). However, we excluded any neoplasm that occurred before the age of 15 years for all patients to attempt to minimize this confounder.

Myelodysplastic syndromes arise from impaired hematopoietic differentiation (30) and are a risk factor for progression to leukemia. One case report detailing an adult with DS described myelodysplastic syndromes as a rare occurrence in the adult DS population (31). However, we observed that individuals aged 40 years and older with DS were more likely to be diagnosed with myelodysplastic syndromes compared to individuals without DS, highlighting the power of our large sample size to identify more rare hematological conditions. Myelodysplastic syndromes are a risk factor for hematological malignancies, but because data are reported in aggregate, we were unable to discern the degree to which myelodysplastic syndromes precede the development of hematological malignancies in our study.

Adults ages ≥40 and older with DS had a higher incidence of multiple myeloma compared to those without DS. Hasle et al. found no multiple myeloma cases in their study (14). However, our study consisted of a larger group of individuals with DS and sufficient power to quantify rare malignancies. A single case report on multiple myeloma in an adult with DS identified it as a rare incident with no established treatment protocols and treatment complications (32). Given that multiple myeloma is associated with aging, with a median age of 69 at diagnosis in the general population, our data suggest that the DS population is gradually developing previously underappreciated age-related malignancies (33). Therefore, creating clearer therapeutic guidelines for multiple myeloma in the adult DS population may be necessary. More studies are needed to determine the factors increasing the predisposition of adults with DS to these hematological malignancies, including studies of clonal hematopoiesis and obesity as a potential risk factor in DS where the incidence is higher (34). That said, while obesity is a risk factor for multiple myeloma, obese patients are better able to tolerate aggressive treatment regimens (35). It is not known whether this translates to DS. It is also unknown if the driving factor for the increased incidence of myeloma is a higher rate of monoclonal gammopathy of undetermined significance (MGUS), a pre-malignant condition, as observed in individuals of African descent, or an increased rate of progression from MGUS to multiple myeloma (36). Future studies should focus on addressing these unknowns in DS, including guidelines to aid in the prevention and early detection of hematological malignancies.

### Solid tumors

Consistent with previous reports, we found a similar prevalence of many malignancies and a lower prevalence of certain solid tumors in our matched cohorts (11, 13, 14). Upon classifying these malignancies into specific subgroups, we observed a higher incidence of testicular cancer in adults (≥ 40 years old) with DS. We found a similar incidence of testicular cancer in AYA with and without DS. Combining all age groups, others also observed an increased incidence of testicular cancer (11, 14).

Individuals with DS have a high incidence of cryptorchidism, which is a risk factor for seminoma, a subtype of testicular cancer (37). The authors of a case report on seminoma in an adult with DS recommended that individuals with DS aged 15 to 45 undergo annual testicular examinations (38). The ≥40-year-old cohort with DS had a significantly lower incidence of breast and prostate cancer, which was persistent when both groups were combined, and a lower incidence of skin and cervical cancer when the groups were combined.

The lower occurrence of some solid tumors in the DS population could be due to several factors, including the gene dosage of tumor suppressor genes on human chromosome 21, increased apoptosis, and DNA damage associated with DS (40). The reduced incidence of solid tumors in DS may be attributed to the inhibition of VEGF-mediated angiogenesis due to the overexpression of *DSCR1* and *DYRK1a* on human chromosome 21 (41). Environmental factors likely also play a role. Compared to the cohort without DS, individuals with DS had less documented exposure to environmental risk factors for cancer development, such as alcohol-related disorders and tobacco use. While the degree to which this is consistently documented in the medical record is not clear, it offers an intriguing environmental hypothesis for cancer risk. Recently, alcohol use has been documented as an important risk factor for breast and prostate cancer (42). It has been suggested that women with DS experience early menopause, which may reduce their risk of breast cancer (39). It is essential to note that while adults with DS may have a lower incidence of certain solid tumors, they are not protected from these conditions. Hence, regular screening remains necessary. Additionally, they share a similar risk for some cancers and should undergo the same routine screenings as the general population.

### Cardiotoxicity

Anthracycline-based regimens are broadly used to treat several cancer types (19, 43). Some underlying risk factors of cardiotoxicity in pediatric patients after anthracycline exposure include age, type of treatment, sex, genetic factors such as DS, cumulative dose, and pre-existing cardiovascular disease (18, 44). After observing a higher incidence of some clinical manifestations of anthracycline cardiotoxicity, such as heart failure and ischemic heart diseases, we explored the DS population to investigate whether our observations were due to our previously documented increased overall risk of cardiovascular disease in DS (21) [Supplemental Table 6]. We found that even within the DS population, there was still an increased risk of cardiovascular disease with anthracycline treatment.

Our findings in the DS population were compared to those of individuals without DS, where we also found increased risk [Supplemental Table 8]. Therefore, even though anthracyclines lead to cardiovascular toxicity generally, this risk is increased in adults with DS and may be enhanced by their inherent cardiovascular disease risk. The increased incidence of cardiovascular diseases in our research serves as a basis to further mechanistically investigate cardiovascular complications in adults with DS.

Finally, as noted in our earlier study, individuals with DS experience a higher incidence of cardiovascular diseases compared to age- and sex-matched peers without DS (21). This suggests that pre-existing increased cardiovascular risk in DS may exacerbate the effects of cardiotoxic therapy in this population. Patients with DS were more likely to be hypotensive and hypertensive prior to cancer diagnoses. This emphasizes the necessity for mechanistically driven studies and organized and individualized care for this unique population.

### Strengths and Limitations

An essential feature of this study is the large cohort of adults with DS that allows for increased power to detect rarer cancers, including multiple myeloma. However, the limitations of this study include our inability to access individual patient records and perform case-control matching based on the date of diagnosis. Due to the retrospective nature of this study, the data shows association and not causality. Using ICD-10 codes is subject to miscoding and non-specific diagnoses. We also do not know how many patients had multiple neoplasm diagnoses, so we did not correct for possible multiple comparisons. Lastly, some healthcare centers for individuals with disabilities may not be part of the TriNetX network, resulting in missed data from those centers, although our data set represents a diverse geographic area and includes most of the large centers that provide comprehensive cancer and developmental care. Because patients cannot be compared between geographic regions, we do not know whether risk differences are geographically driven.

## CONCLUSIONS

We found that individuals with DS are more likely to develop cardiovascular diseases after treatment with anthracyclines and more likely to develop age-related hematological malignancies than previously reported. Diseases like multiple myeloma, considered rare in individuals with DS, should be reexamined as life expectancy continues to rise. Despite the low incidence of solid tumors in this group, they are not entirely protected from these malignancies and are at similar risk for some. It is essential to consider appropriate screening methods and timing for adults with DS as they age. Given the differences between our matched and unmatched cohorts for some cancers, DS individuals may experience these tumors earlier than typically seen in the general population. The absence of established healthcare guidelines for AYA and adult DS patients may result in lower survival rates, potentially exclude them from important clinical studies, and reduce life expectancy. Given the increased risk of cardiovascular disease in DS individuals, increased screening following cardiotoxic chemotherapy may be warranted. Studies examining clonal hematopoiesis and the effects of aging on bone marrow function in adults with DS are warranted. Our limited understanding, along with the results from this extensive real-world cohort study, underlines the need for future cancer screening and survivorship guidelines to be tailored to the specific needs of the aging DS population.

## Author Contribution

MAB, AV, CRJ, HK, MV, MHT, MLB – Participated in the Conceptualization, method design, data collection and analysis, interpretation of results, figure and manuscript preparation, and gave approval of the final version.

## Grants or Funding

This work was supported by the National Institute of Health R01CA244271 (Bates and Tomasson) and American Cancer Society RSG-20-017-01-CCE (Bates and Tomasson).

## Data Availability

All data produced in the present study are available upon reasonable request to the authors

---

**Supplemental Table 1.**
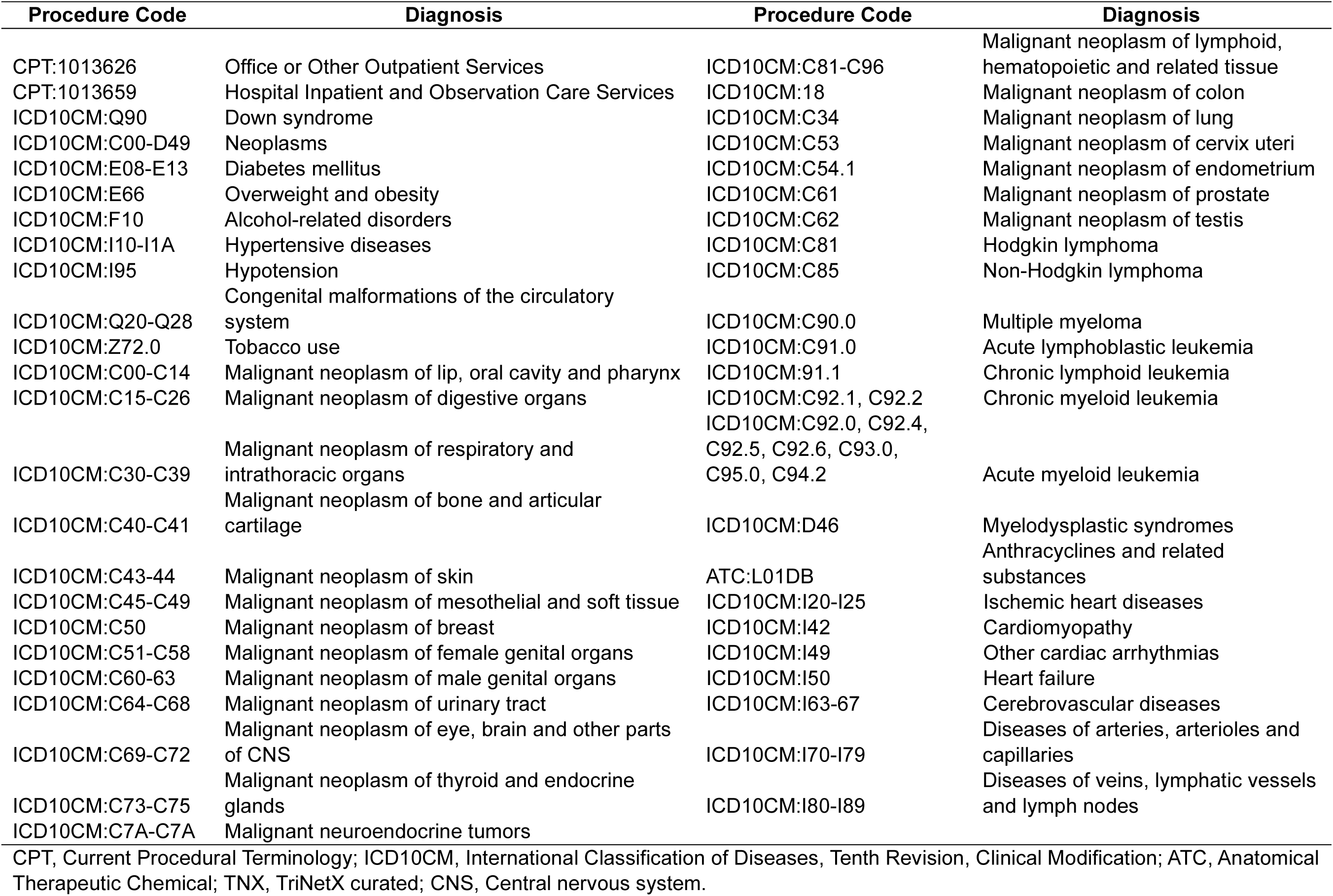
Diagnosis and procedure codes used for TriNetX search.

**Supplemental Table 2.**
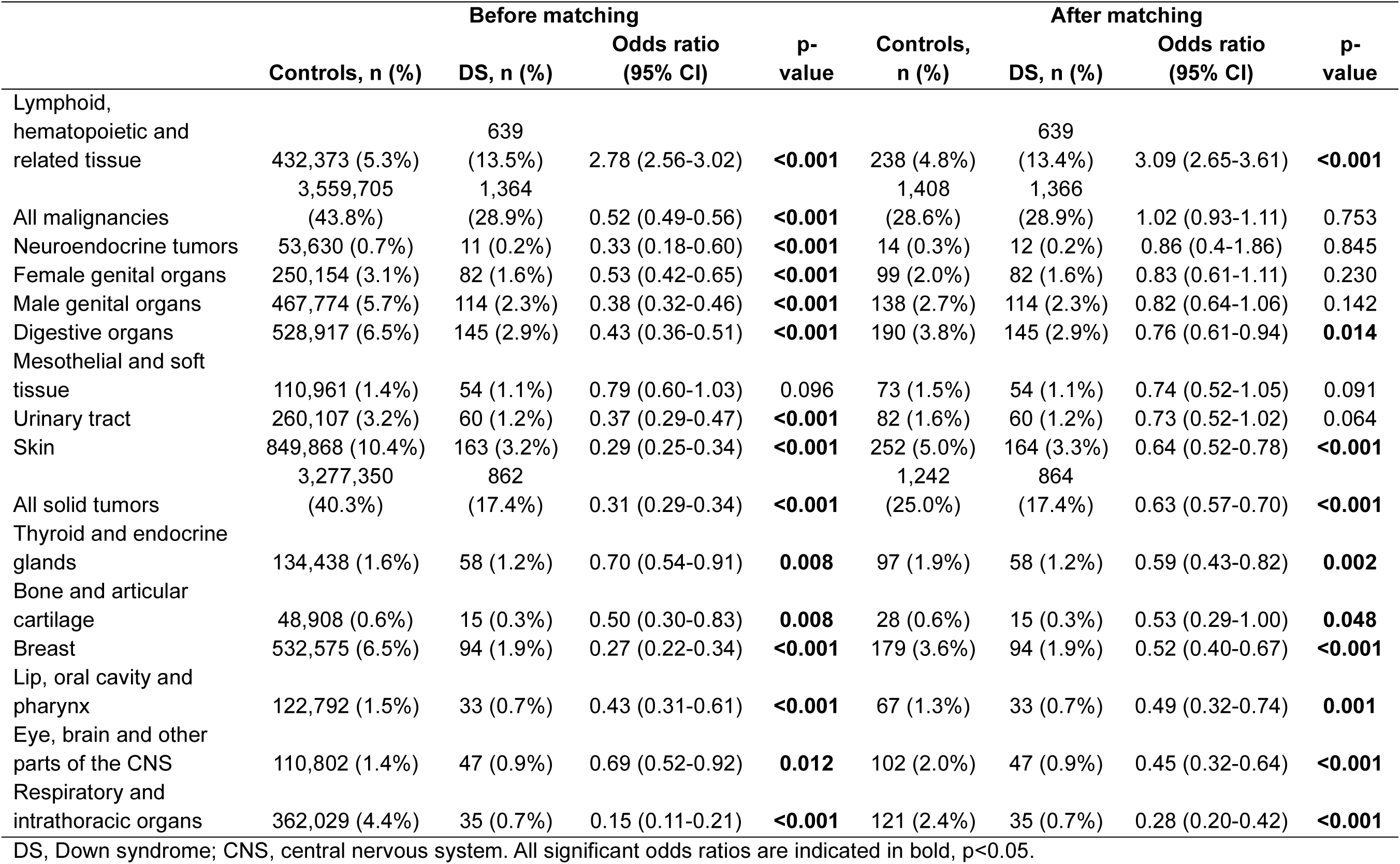
The incidence of cancer in the major body sites before and after matching.

**Supplemental Table 3.** Baseline characteristics of patients with and without DS after treatment with anthracyclines.

|  | Before matching |  |  | After matching |  |  |
| --- | --- | --- | --- | --- | --- | --- |
|  | Controls<br>n=102,976 | DS<br>n=177 | p-value | Controls<br>n=175 | DS<br>n=175 | p-value |
| **Age, years** |  |  |  |  |  |  |
| †Age at index | 55.2 ± 15.9 | 41.7 ± 18.6 | **< 0.001** | 41.7 ± 18.6 | 41.7 ± 18.6 | 1.000 |
| Current age | 62.1 ± 16.1 | 48.6 ± 18.9 | **< 0.001** | 48.7 ± 19.3 | 48.6 ± 18.9 | 0.940 |
| **Sex** |  |  |  |  |  |  |
| †Female | 58,704 (58.8%) | 77 (44%) | **< 0.001** | 77 (44.0%) | 77 (44.0%) | 1.000 |
| Male | 38,025 (38.1%) | 94 (53.7%) | **< 0.001** | 92 (52.6%) | 94 (53.7%) | 0.830 |
| Unknown sex | 3,064 (3.1%) | 10 (5.7%) | **0.043** | 10 (5.7%) | 10 (5.7%) | 1.000 |
| **Race** |  |  |  |  |  |  |
| White | 68,302 (68.4%) | 115 (65.7%) | 0.438 | 120 (68.6%) | 115 (65.7%) | 0.569 |
| Unknown race | 10,621 (10.6%) | 13 (7.4%) | 0.168 | 22 (12.6%) | 13 (7.4%) | 0.109 |
| Black or African American | 11,841 (11.9%) | 19 (10.9%) | 0.680 | 16 (9.1%) | 19 (10.9%) | 0.593 |
| Other Race | 3,751 (3.8%) | 17 (9.7%) | **< 0.001** | 10 (5.7%) | 17 (9.7%) | 0.161 |
| Asian | 4,344 (4.4%) | 10 (5.7%) | 0.378 | 11 (6.3%) | 10 (5.7%) | 0.822 |
| Native Hawaiian or other Pacific Islander | 621 (0.6%) | 10 (5.7%) | **< 0.001** | 0 | 10 (5.7%) | **0.001** |
| American Indian or Alaska Native | 313 (0.3%) | 10 (5.7%) | **< 0.001** | 0 | 10 (5.7%) | **0.001** |
| **Ethnicity** |  |  |  |  |  |  |
| Not Hispanic or Latino | 72,356 (72.5%) | 118 (67.4%) | 0.133 | 118 (67.4%) | 118 (67.4%) | 1.000 |
| Hispanic or Latino | 8,309 (8.3%) | 35 (20.0%) | **< 0.001** | 19 (10.9%) | 35 (20.0%) | **0.018** |
| Unknown Ethnicity | 19,128 (19.2%) | 22 (12.6%) | **0.027** | 38 (21.7%) | 22 (12.6%) | **0.023** |
| **Comorbidities and lifestyle factors** |  |  |  |  |  |  |
| Congenital malformations of the circulatory system | 1,856 (1.9%) | 23 (13.1%) | **< 0.001** | 10 (5.7%) | 23 (13.1%) | **0.017** |
| Diabetes mellitus | 17,839 (17.9%) | 33 (18.9%) | 0.735 | 23 (13.1%) | 33 (18.9%) | 0.145 |
| Hypotension | 9,167 (9.2%) | 25 (14.3%) | **0.020** | 22 (12.6%) | 25 (14.3%) | 0.638 |
| Hypertensive diseases | 42,174 (42.3%) | 66 (37.7%) | 0.224 | 47 (26.9%) | 66 (37.7%) | **0.030** |
| Overweight and obesity | 19,736 (19.8%) | 45 (25.7%) | 0.049 | 28 (16.0%) | 45 (25.7%) | **0.025** |

Data are represented as mean ± SD or n (%). DS, Down syndrome. All significant values are indicated in bold, p<0.05. †Demographic parameters used to match groups.

**Supplemental Table 4.** The incidence of cardiovascular diseases in adults with and without DS after treatment with anthracyclines.

|  | Before matching |  |  |  | After matching |  |  |  |
| --- | --- | --- | --- | --- | --- | --- | --- | --- |
|  | Controls, n (%) | DS, n (%) | Odds ratio<br>(95% CI) | p-value | Controls,<br>n (%) | DS, n (%) | Odds ratio<br>(95% CI) | p-value |
| Ischemic heart diseases | 11,988 (13.7%) | 21 (13.1%) | 0.95 (0.60-1.50) | 0.909 | 10 (6.2%) | 21 (13.1%) | 2.30 (1.05-5.05) | **0.039** |
| Diseases of arteries, arterioles and capillaries | 11,366 (12.8%) | 28 (17.7%) | 1.47 (0.98-2.22) | 0.073 | 14 (8.6%) | 28 (17.7%) | 2.28 (1.15-4.51) | **0.020** |
| Heart failure | 10,195 (10.9%) | 26 (16.6%) | 1.63 (1.07-2.48) | **0.029** | 14 (8.5%) | 26 (16.6%) | 2.14 (1.07-4.27) | **0.042** |
| Cerebrovascular diseases | 6,811 (7.2%) | 19 (11.3%) | 1.64 (1.02-2.65) | 0.050 | 10 (5.8%) | 19 (11.3%) | 2.07 (0.93-4.59) | 0.082 |
| Diseases of veins, lymphatic vessels and lymph nodes | 20,892 (26.2%) | 35 (25.9%) | 0.98 (0.67-1.45) | 1.000 | 22 (15.9%) | 35 (25.9%) | 1.85 (1.02-3.35) | 0.053 |
| Cardiomyopathy | 6,716 (7.0%) | 18 (10.8%) | 1.61 (0.99-2.63) | 0.066 | 14 (8.2%) | 18 (10.8%) | 1.36 (0.65-2.82) | 0.461 |
| Other cardiac arrhythmias | 12,482 (13.9%) | 21 (14.1%) | 1.01 (0.64-1.61) | 0.906 | 27 (17.2%) | 21 (14.1%) | 0.79 (0.43-1.47) | 0.530 |

DS, Down syndrome. All significant values are indicated in bold, $p < 0.05$ .

**Supplemental Table 5.** Baseline characteristics with Down syndrome.

|  | Before matching |  |  | After matching |  |  |
| --- | --- | --- | --- | --- | --- | --- |
|  | No<br>anthracyclines<br>n=35,229 | Anthracyclines<br>n=177 | p-value | DS-no<br>anthracyclines<br>n=175 | DS<br>n=175 | p-value |
| **Age, years** |  |  |  |  |  |  |
| †Age at index | 24.1 ± 18.0 | 41.7 ± 18.6 | **< 0.001** | 41.7 ± 18.6 | 41.7 ± 18.6 | 1.000 |
| Current age | 33.8 ± 16.5 | 48.6 ± 18.9 | **< 0.001** | 50.3 ± 19.3 | 48.6 ± 18.9 | 0.393 |
| **Sex** |  |  |  |  |  |  |
| †Female | 15,332 (48.9%) | 77 (44.0%) | 0.192 | 77 (44.0%) | 77 (44.0%) | 1.000 |
| Male | 15,283 (48.8%) | 94 (53.7%) | 0.193 | 92 (52.6%) | 94 (53.7%) | 0.830 |
| Unknown sex | 713 (2.3%) | 10 (5.7%) | **0.003** | 10 (5.7%) | 10 (5.7%) | 1.000 |
| **Race** |  |  |  |  |  |  |
| White | 20,783 (66.3%) | 115 (65.7%) | 0.861 | 119 (68.0%) | 115 (65.7%) | 0.650 |
| Unknown race | 4,745 (15.1%) | 13 (7.4%) | **0.005** | 25 (14.3%) | 13 (7.4%) | **0.039** |
| Black or African American | 3,056 (9.8%) | 19 (10.9%) | 0.624 | 12 (6.9%) | 19 (10.9%) | 0.188 |
| Other Race | 1,758 (5.6%) | 17 (9.7%) | **0.019** | 12 (6.9%) | 17 (9.7%) | 0.332 |
| Asian | 828 (2.6%) | 10 (5.7%) | **0.012** | 10 (5.7%) | 10 (5.7%) | 1.000 |
| Native Hawaiian or other Pacific Islander | 70 (0.2%) | 10 (5.7%) | **< 0.001** | 10 (5.7%) | 10 (5.7%) | 1.000 |
| American Indian or Alaska Native | 88 (0.3%) | 10 (5.7%) | **< 0.001** | 0 | 10 (5.7%) | **0.001** |
| **Ethnicity** |  |  |  |  |  |  |
| Not Hispanic or Latino | 19,826 (63.3%) | 118 (67.4%) | 0.257 | 108 (61.7%) | 118 (67.4%) | 0.264 |
| Hispanic or Latino | 5,060 (16.2%) | 35 (20.0%) | 0.168 | 24 (13.7%) | 35 (20.0%) | 0.116 |
| Unknown Ethnicity | 6,442 (20.6%) | 22 (12.6%) | **0.009** | 43 (24.6%) | 22 (12.6%) | **0.004** |
| **Comorbidities and lifestyle factors** |  |  |  |  |  |  |
| Congenital malformations of the circulatory system | 2,280 (7.3%) | 23 (13.1%) | **0.003** | 10 (5.7%) | 23 (13.1%) | **0.017** |
| Diabetes mellitus | 725 (2.3%) | 33 (18.9%) | **< 0.001** | 10 (5.7%) | 33 (18.9%) | **< 0.001** |
| Hypotension | 237 (0.8%) | 25 (14.3%) | **< 0.001** | 10 (5.7%) | 25 (14.3%) | **0.008** |
| Hypertensive diseases | 726 (2.3%) | 66 (37.7%) | **< 0.001** | 10 (5.7%) | 66 (37.7%) | **< 0.001** |
| Overweight and obesity | 1,033 (3.3%) | 45 (25.7%) | **< 0.001** | 10 (5.7%) | 45 (25.7%) | **< 0.001** |

Data are represented as mean ± SD or n (%). DS, Down syndrome. All significant values are indicated in bold, p<0.05. †Demographic parameters used to match groups.

**Supplemental Table 6.** The incidence of cardiovascular diseases in adults with Down syndrome comparing those with and without anthracyclines.

|  | Before matching |  |  |  | After matching |  |  |  |
| --- | --- | --- | --- | --- | --- | --- | --- | --- |
|  | No anthracyclines, n (%) | Anthracyclines, n (%) | Odds ratio (95% CI) | p-value | No anthracyclines, n (%) | Anthracyclines, n (%) | Odds ratio (95% CI) | p-value |
| Ischemic heart diseases | 605 (2.0%) | 21 (13.1%) | 7.60 (4.77-12.1) | **<0.001** | 10 (6.0%) | 21 (13.1%) | 2.36 (1.07-5.18) | **0.037** |
| Diseases of arteries, arterioles and capillaries | 918 (3.0%) | 28 (17.7%) | 7.06 (4.67-10.67) | **<0.001** | 10 (5.9%) | 28 (17.7%) | 3.45 (1.61-7.36) | **0.001** |
| Heart failure | 1,113 (3.6%) | 26 (16.6%) | 5.28 (3.45-8.07) | **<0.001** | 10 (6.1%) | 26 (16.6%) | 3.08 (1.43-6.61) | **0.004** |
| Cerebrovascular diseases | 658 (2.1%) | 19 (11.3%) | 5.85 (3.60-9.48) | **<0.001** | 10 (6.0%) | 19 (11.3%) | 2.02 (0.91-4.47) | 0.119 |
| Diseases of veins, lymphatic vessels and lymph nodes | 1,286 (4.2%) | 35 (25.9%) | 8.01 (5.43-11.82) | **<0.001** | 10 (6.0%) | 35 (25.9%) | 5.50 (2.61-11.59) | **<0.001** |
| Cardiomyopathy | 316 (1.0%) | 18 (10.8%) | 11.87 (7.19-19.61) | **<0.001** | 10 (5.9%) | 18 (10.8%) | 1.95 (0.87-4.35) | 0.116 |
| Other cardiac arrhythmias | 1,998 (6.6%) | 21 (14.1%) | 2.32 (1.46-3.69) | **0.001** | 10 (5.8%) | 21 (14.1%) | 2.67 (1.22-5.88) | **0.014** |

DS, Down syndrome. All significant values are indicated in bold, p<0.05.

**Supplemental Table 7.** Baseline characteristics of patients without DS.

|  | Before matching |  |  | After matching |  |  |
| --- | --- | --- | --- | --- | --- | --- |
|  | No Ds and anthracyclines<br>n=31,145,476 | No DS and no anthracyclines<br>n=102,976 | p-value | Controls<br>n=99,790 | DS<br>n=99,790 | p-value |
| **Age, years** |  |  |  |  |  |  |
| †Age at index | 40.6 ± 22.1 | 55.2 ± 15.9 | **< 0.001** | 55.2 ± 15.9 | 55.2 ± 15.9 | 1.000 |
| Current age | 47.7 ± 21.5 | 62.1 ± 16.1 | **< 0.001** | 61.9 ± 16.3 | 62.1 ± 16.1 | 0.090 |
| **Sex** |  |  |  |  |  |  |
| †Female | 15,730,104 (52.7%) | 58,704 (58.8%) | **< 0.001** | 58,704 (58.8%) | 58,704 (58.8%) | 1.000 |
| Male | 13,135,405 (44.0%) | 38,022 (38.1%) | **< 0.001** | 37,931 (38.0%) | 38,022 (38.1%) | 0.675 |
| Unknown sex | 955,744 (3.2%) | 3,064 (3.1%) | **0.016** | 3,155 (3.2%) | 3,064 (3.1%) | 0.241 |
| **Race** |  |  |  |  |  |  |
| White | 17,857,203 (59.9%) | 68,299 (68.4%) | **< 0.001** | 62,327 (62.5%) | 68,299 (68.4%) | **< 0.001** |
| Unknown race | 5,535,554 (18.6%) | 10,621 (10.6%) | **< 0.001** | 18,080 (18.1%) | 10,621 (10.6%) | **< 0.001** |
| Black or African American | 3,647,348 (12.2%) | 11,841 (11.9%) | **< 0.001** | 11,103 (11.1%) | 11,841 (11.9%) | **< 0.001** |
| Other Race | 1,547,991 (5.2%) | 3,751 (3.8%) | **< 0.001** | 4,348 (4.4%) | 3,751 (3.8%) | **< 0.001** |
| Asian | 1,043,983 (3.5%) | 4,344 (4.4%) | **< 0.001** | 3,362 (3.4%) | 4,344 (4.4%) | **< 0.001** |
| Native Hawaiian or other Pacific Islander | 105,139 (0.4%) | 621 (0.6%) | **< 0.001** | 341 (0.3%) | 621 (0.6%) | **< 0.001** |
| American Indian or Alaska Native | 84,035 (0.3%) | 313 (0.3%) | 0.058 | 229 (0.2%) | 313 (0.3%) | **< 0.001** |
| **Ethnicity** |  |  |  |  |  |  |
| Not Hispanic or Latino | 18,756,726 (62.9%) | 72,354 (72.5%) | **< 0.001** | 64,669 (64.8%) | 72,354 (72.5%) | **< 0.001** |
| Hispanic or Latino | 3,412,931 (11.4%) | 8,309 (8.3%) | **< 0.001** | 8,528 (8.5%) | 8,309 (8.3%) | 0.078 |
| Unknown Ethnicity | 7,651,596 (25.7%) | 19,127 (19.2%) | **< 0.001** | 26,593 (26.6%) | 19,127 (19.2%) | **< 0.001** |
| **Comorbidities and lifestyle factors** |  |  |  |  |  |  |
| Congenital malformations of the circulatory system | 135,883 (0.5%) | 1,856 (1.9%) | **< 0.001** | 410 (0.4%) | 1,856 (1.9%) | **< 0.001** |
| Diabetes mellitus | 1,140,034 (3.8%) | 17,839 (17.9%) | **< 0.001** | 5,981 (6.0%) | 17,839 (17.9%) | **< 0.001** |
| Hypotension | 205,306 (0.7%) | 9,167 (9.2%) | **< 0.001** | 978 (1.0%) | 9,167 (9.2%) | **< 0.001** |
| Hypertensive diseases | 2,478,832 (8.3%) | 42,174 (42.3%) | **< 0.001** | 13,024 (13.1%) | 42,174 (42.3%) | **< 0.001** |
| Overweight and obesity | 1,069,165 (3.6%) | 19,736 (19.8%) | **< 0.001** | 4,293 (4.3%) | 19,736 (19.8%) | **< 0.001** |

Data are represented as mean ± SD or n (%). DS, Down syndrome. All significant values are indicated in bold, p<0.05. †Demographic parameters used to match groups.

**Supplemental Table 8.** The incidence of cardiovascular diseases in adults without Down syndrome comparing the incidence of cardiovascular disease after treatment with anthracyclines.

|  | Before matching |  |  |  | After matching |  |  |  |
| --- | --- | --- | --- | --- | --- | --- | --- | --- |
|  | No anthracyclines, n (%) | Anthracyclines, n (%) | Odds ratio (95% CI) | p-value | No anthracyclines, n (%) | Anthracyclines, n (%) | Odds ratio (95% CI) | p-value |
| Ischemic heart diseases | 1,160,167 (4.1%) | 11,988 (13.7%) | 3.75 (3.68-3.83) | **<0.001** | 6,424 (6.9%) | 11,988 (13.7%) | 2.14 (2.07-2.20) | **<0.001** |
| Diseases of arteries, arterioles and capillaries | 947,941 (3.3%) | 11,365 (12.8%) | 4.35 (4.27-4.44) | **<0.001** | 5,113 (5.3%) | 11,365 (12.8%) | 2.61 (2.52-2.70) | **<0.001** |
| Heart failure | 730,936 (2.5%) | 10,195 (10.9%) | 4.75 (4.66-4.85) | **<0.001** | 3,876 (4.0%) | 10,195 (10.9%) | 2.92 (2.81-3.03) | **<0.001** |
| Cerebrovascular diseases | 686,193 (2.4%) | 6,810 (7.2%) | 3.22 (3.14-3.30) | **<0.001** | 3,717 (3.9%) | 6,810 (7.2%) | 1.93 (1.86-2.01) | **<0.001** |
| Diseases of veins, lymphatic vessels and lymph nodes | 791,224 (2.7%) | 20,891 (26.2%) | 12.8 (12.60-13.01) | **<0.001** | 3,519 (3.6%) | 20,891 (26.2%) | 9.47 (9.12-9.83) | **<0.001** |
| Cardiomyopathy | 313,835 (1.1%) | 6,716 (7.0%) | 7.04 (6.87-7.22) | **<0.001** | 1,597 (1.6%) | 6,716 (7.0%) | 4.59 (4.34-4.85) | **<0.001** |
| Other cardiac arrhythmias | 1,079,634 (3.7%) | 12,482 (13.9%) | 4.22 (4.14-4.30) | **<0.001** | 4,630 (4.8%) | 12,482 (13.9%) | 3.24 (3.12-3.35) | **<0.001** |

DS, Down syndrome. All significant values are indicated in bold, p<0.05.

## REFERENCES

1. De Graaf G, Buckley F, and Skotko BG. Estimation of the number of people with Down syndrome in the United States. Genetics in Medicine 19: 439–447, 2017.

2. Presson AP, Partyka G, Jensen KM, Devine OJ, Rasmussen SA, McCabe LL, and McCabe ER. Current estimate of Down Syndrome population prevalence in the United States. J Pediatr 163: 1163–1168, 2013.

3. Antonarakis SE, Skotko BG, Rafii MS, Strydom A, Pape SE, Bianchi DW, Sherman SL, and Reeves RH. Down syndrome. Nature Reviews Disease Primers 6: 2020.

4. Bull MJ. Health Supervision for Children With Down Syndrome. Pediatrics 128: 393–406, 2011.

5. Zigman WB. Atypical aging in Down syndrome. Dev Disabil Res Rev 18: 51–67, 2013.

6. Muchová J, S̆ustrová M, Garaiová I, Liptáková A, Blaz̆íc̆ek P, Kvasnic̆ka P, Pueschel S, and D̆urac̆ková Zk. Influence of age on activities of antioxidant enzymes and lipid peroxidation products in erythrocytes and neutrophils of Down syndrome patients. Free Radical Biology and Medicine 31: 499–508, 2001.

7. Helguera P, Pelsman A, Pigino G, Wolvetang E, Head E, and Busciglio J. ets-2 Promotes the Activation of a Mitochondrial Death Pathway in Down’s Syndrome Neurons. The Journal of Neuroscience 25: 2295–2303, 2005.

8. Esposito G, Imitola J, Lu J, De Filippis D, Scuderi C, Ganesh VS, Folkerth R, Hecht J, Shin S, Iuvone T, Chesnut J, Steardo L, and Sheen V. Genomic and functional profiling of human Down syndrome neural progenitors implicates S100B and aquaporin 4 in cell injury. Human Molecular Genetics 17: 440–457, 2008.

9. Izzo A, Manco R, Bonfiglio F, Calì G, De Cristofaro T, Patergnani S, Cicatiello R, Scrima R, Zannini M, Pinton P, Conti A, and Nitsch L. NRIP1/RIP140 siRNA-mediated attenuation counteracts mitochondrial dysfunction in Down syndrome. Human Molecular Genetics 23: 4406–4419, 2014.

10. Hasle H, Clemmensen IH, and Mikkelsen M. Risks of leukaemia and solid tumours in individuals with Down’s syndrome. Lancet 355: 165–169, 2000.

11. Patja K, Pukkala E, Sund R, Iivanainen M, and Kaski M. Cancer incidence of persons with down syndrome in Finland: A population-based study. International Journal of Cancer 118: 1769–1772, 2006.

12. Hill DA, Gridley G, Cnattingius S, Mellemkjaer L, Linet M, Adami H-O, Olsen JH, Nyren O, and Fraumeni JF. Mortality and Cancer Incidence Among Individuals With Down Syndrome. Archives of Internal Medicine 163: 705, 2003.

13. Krieg S, Krieg A, Loosen SH, Roderburg C, and Kostev K. Cancer Risk in Patients with Down Syndrome—A Retrospective Cohort Study from Germany. Cancers 16: 1103, 2024.

14. Hasle H, Friedman JM, Olsen JH, and Rasmussen SA. Low risk of solid tumors in persons with Down syndrome. Genetics in Medicine 18: 1151–1157, 2016.

15. Rethore MO, Rouesse J, and Satge D. Cancer screening in adults with down syndrome, a proposal. Eur J Med Genet 63: 103783, 2020.

16. Tsou AY, Bulova P, Capone G, Chicoine B, Gelaro B, Harville TO, Martin BA, McGuire DE, McKelvey KD, Peterson M, Tyler C, Wells M, and Whitten MS. Medical Care of Adults With Down Syndrome. JAMA 324: 1543, 2020.

17. Henriksen PA. Anthracycline cardiotoxicity: an update on mechanisms, monitoring and prevention. Heart 104: 971–977, 2018.

18. Krischer JP, Epstein S, Cuthbertson DD, Goorin AM, Epstein ML, and Lipshultz SE. Clinical cardiotoxicity following anthracycline treatment for childhood cancer: the Pediatric Oncology Group experience. J Clin Oncol 15: 1544–1552, 1997.

19. Hefti E, and Blanco JG. Anthracycline-Related Cardiotoxicity in Patients with Acute Myeloid Leukemia and Down Syndrome: A Literature Review. Cardiovascular Toxicology 16: 5–13, 2016.

20. Zamorano JL, Lancellotti P, Rodriguez Muñoz D, Aboyans V, Asteggiano R, Galderisi M, Habib G, Lenihan DJ, Lip GYH, Lyon AR, Lopez Fernandez T, Mohty D, Piepoli MF, Tamargo J, Torbicki A, and Suter TM. 2016 ESC Position Paper on cancer treatments and cardiovascular toxicity developed under the auspices of the ESC Committee for Practice Guidelines. European Heart Journal 37: 2768–2801, 2016.

21. Bates ML, Vasileva A, Flores LDM, Pryakhina Y, Buckman M, Tomasson MH, and DeRuisseau LR. Sex diferences in cardiovascular disease and dysregulation in Down syndrome. Am J Physiol Heart Circ Physiol 324: H542–H552, 2023.

22. Janardan SK, and Miller TP. Adolescents and young adults (AYAs) vs pediatric patients: survival, risks, and barriers to enrollment. Hematology 2023: 581–586, 2023.

23. Deruisseau LR, Receno CN, Cunningham C, Bates ML, Goodell M, Liang C, Eassa B, Pascolla J, and Deruisseau KC. Breathing and Oxygen Carrying Capacity in Ts65Dn and Down Syndrome. Function 4: 2023.

24. Baruchel A, Bourquin J-P, Crispino J, Cuartero S, Hasle H, Hitzler J, Klusmann J-H, Izraeli S, Lane AA, Malinge S, Rabin KR, Roberts I, Ryeom S, Tasian SK, and Wagenblast E. Down syndrome and leukemia: from basic mechanisms to clinical advances. Haematologica 108: 2570–2581, 2023.

25. De Castro CPM, Cadefau M, and Cuartero S. The Mutational Landscape of Myeloid Leukaemia in Down Syndrome. Cancers 13: 4144, 2021.

26. Zaslavsky A, Chou ST, Schadler K, Lieberman A, Pimkin M, Kim YJ, Baek K-H, Aird WC, Weiss MJ, and Ryeom S. The calcineurin-NFAT pathway negatively regulates megakaryopoiesis. Blood 121: 3205–3215, 2013.

27. Rainis L, Toki T, Pimanda JE, Rosenthal E, Machol K, Strehl S, GöTtgens B, Ito E, and Izraeli S. The Proto-Oncogene ERG in Megakaryoblastic Leukemias. Cancer Research 65: 7596–7602, 2005.

28. Cabal-Hierro L, Van Galen P, Prado MA, Higby KJ, Togami K, Mowery CT, Paulo JA, Xie Y, Cejas P, Furusawa T, Bustin M, Long HW, Sykes DB, Gygi SP, Finley D, Bernstein BE, and Lane AA. Chromatin accessibility promotes hematopoietic and leukemia stem cell activity. Nature Communications 11: 2020.

29. Michels N, Boer JM, Enshaei A, Sutton R, Heyman M, Ebert S, Fiocco M, De Groot-Kruseman HA, Van Der Velden VHJ, Barbany G, Escherich G, Vora A, Trahair T, Dalla-Pozza L, Pieters R, Zur Stadt U, Schmiegelow K, Moorman AV, Zwaan CM, and Den Boer ML. Minimal residual disease, long-term outcome, and IKZF1 deletions in children and adolescents with Down syndrome and acute lymphocytic leukaemia: a matched cohort study. The Lancet Haematology 8: e700–e710, 2021.

30. Hitzler JK, and Zipursky A. Origins of leukaemia in children with Down syndrome. Nature Reviews Cancer 5: 11–20, 2005.

31. Virk H, and Naseem S. A Case of Myelodysplastic Syndrome in an Adult with Down Syndrome: A Rare Observation of a Well-known Pediatric Disease. Turk J Haematol 37: 137–138, 2020.

32. Musleh M, and Alhusein Q. A rare presentation and treatment challenges for multiple myeloma in down syndrome: A case report and literature review. Clinical Case Reports 12: 2024.

33. Greteman BB, Tomasson MH, Kahl AR, Wahlen MM, Bates ML, Strouse C, and Charlton ME. Comparison of outcomes by race among a population-based matched sample of multiple myeloma patients. Cancer Causes & Control 2024.

34. Carson KR, Bates ML, and Tomasson MH. The skinny on obesity and plasma cell myeloma: a review of the literature. Bone Marrow Transplantation 49: 1009–1015, 2014.

35. Beason TS, Chang S-H, Sanfilippo KM, Luo S, Colditz GA, Vij R, Tomasson MH, Dipersio JF, Stockerl-Goldstein K, Ganti A, Wildes T, and Carson KR. Influence of Body Mass Index on Survival in Veterans With Multiple Myeloma. The Oncologist 18: 1074–1079, 2013.

36. Tomasson MH, Ali M, De Oliveira V, Xiao Q, Jethava Y, Zhan F, Fitzsimmons AM, and Bates ML. Prevention Is the Best Treatment: The Case for Understanding the Transition from Monoclonal Gammopathy of Undetermined Significance to Myeloma. International Journal of Molecular Sciences 19: 3621, 2018.

37. Benson RC, Jr., Beard CM, Kelalis PP, and Kurland LT. Malignant potential of the cryptorchid testis. Mayo Clin Proc 66: 372–378, 1991.

38. Hengy M, Hinge A, Purtell JP, Shango K, and Collins J. Metastatic Testicular Seminoma in a Patient With Down Syndrome Presenting As Extensive Deep Venous Thrombosis. Cureus 2022.

39. Seltzer G, Schupf N, and Wu HS. A prospective study of menopause in women with Down’s syndrome. Journal of Intellectual Disability Research 45: 1–7, 2001.

40. Osuna-Marco MP, López-Barahona M, López-Ibor B, and Tejera ÁM. Ten Reasons Why People With Down Syndrome are Protected From the Development of Most Solid Tumors -A Review. Frontiers in Genetics 12: 2021.

41. Baek KH, Zaslavsky A, Lynch RC, Britt C, Okada Y, Siarey RJ, Lensch MW, Park IH, Yoon SS, Minami T, Korenberg JR, Folkman J, Daley GQ, Aird WC, Galdzicki Z, and Ryeom S. Down’s syndrome suppression of tumour growth and the role of the calcineurin inhibitor DSCR1. Nature 459: 1126–1130, 2009.

42. Teissedre P-L, Rasines-Perea Z, Ruf J-C, Stockley C, Antoce AO, Romano R, Fradera U, and Kosti RI. Efects of alcohol consumption in general, and wine in particular, on the risk of cancer development: a review. OENO One 54: 813–832, 2020.

43. O’Brien MM, Taub JW, Chang MN, Massey GV, Stine KC, Raimondi SC, Becton D, Ravindranath Y, Dahl GV, and Children’s Oncology Group Study POG. Cardiomyopathy in children with Down syndrome treated for acute myeloid leukemia: a report from the Children’s Oncology Group Study POG 9421. J Clin Oncol 26: 414-420, 2008.

44. Venegas-Zamora L, Bravo-Acuna F, Sigcho F, Gomez W, Bustamante-Salazar J, Pedrozo Z, and Parra V. New Molecular and Organelle Alterations Linked to Down Syndrome Heart Disease. Front Genet 12: 792231, 2021.

